# Virtual reality mediated brain-computer interface training improves sensorimotor neuromodulation in unimpaired and post spinal cord injury individuals

**DOI:** 10.1101/2024.12.18.24317160

**Authors:** Malik Muhammad Naeem Mannan, Dinesh B. Palipana, Kyle Mulholland, Evan Jurd, Ewan C.R. Lloyd, Alastair R. J. Quinn, Claire B. Crossley, Muhammad Fazle Rabbi, David G. Lloyd, Yang D. Teng, Claudio Pizzolato

## Abstract

Real-time brain-computer interfaces (BCIs) that decode electroencephalograms (EEG) during motor imagery (MI) are a powerful adjunct to rehabilitation therapy after neurotrauma. Immersive virtual reality (VR) could complement BCIs by delivering multisensory feedback congruent to the user’s MI, enabling therapies that engage users in task-oriented scenarios. Yet, therapeutic outcomes rely on the user’s proficiency in evoking MI to attain volitional BCI-commanded VR interaction. While previous studies suggested that users could improve BCI-evoked MI within a single session, the effects of multiple training sessions on sensorimotor neuromodulation remain unknown. Here, we present a longitudinal study assessing the impact of VR-mediated BCI training on lower-limb sensorimotor neuromodulation, wherein an EEG-based BCI was coupled with congruent real-time multisensory feedback in immersive VR. We show that unimpaired individuals could learn to modulate their sensorimotor activations during MI virtual walking over multiple training sessions, also resulting in increased BCI control accuracy. Additionally, when extending the system to immersive VR cycling, four individuals with chronic complete spinal cord injury (SCI) showed similar improvements. This is the first study demonstrating that individuals could learn modulating sensorimotor activity associated with MI using BCI integrated with immersive VR over multiple training sessions, even after SCI-induced motor and sensory decline. These results suggest that VR-BCI training may facilitate neuroplasticity, potentially strengthening sensorimotor pathways and functional connectivity relevant to motor control and recovery.

## Introduction

A brain-computer interface (BCI) converts neural signals, such as electroencephalograms (EEGs), into commands that represent specific cerebral information without directly involving the spinal cord, peripheral nervous system, and muscles^1,2^. Motor imagery (MI), which involves imagining a motor task^3^, allows individuals to interact with real-world or virtual environments using BCIs^4,5^. It has been proposed that BCIs can potentiate the effect of sensorimotor rehabilitation after stroke and spinal cord injury (SCI)^5–9^. However, the classification accuracy of BCIs during rehabilitation is limited by the ability of the user to volitionally evoke repeatable EEG patterns^10^. Previous studies suggested that individuals can improve accuracy of BCI-commanded interactions^11^, but investigations have been limited to a small number of training sessions^12–15^. Consequently, the observed improvements may be transitory rather than reflecting consolidated learning. Furthermore, sustaining MI for longer periods—through continuous control—is crucial for effective rehabilitation^13^, as shorter tasks commonly employed in conventional BCI may not adequately support the development of MI control necessary for therapeutic gains, particularly in training involving locomotion tasks^9,13,16^.

Visualizing a limb movement during mirror therapy may engage sensorimotor-related brain networks^17,18^, potentially enhancing motor functions^19^. However, this approach has limited use for dynamic functional tasks, such as walking^20^, and is often compromised by poor user attention and motivation, which may be overcome by using virtual reality (VR)^21^. VR can extend beyond the physical realm conferred by mirror therapy and deliver an immersive and multisensory experience -visual and auditory-that supports cognitive and motor learning^18,22,23^. Furthermore, integrating BCIs with VR enhances user experience in terms of engagement and motivation by leveraging VR game-like environments^21,24^. While previous studies suggest that VR could compliment BCI performance within a single session^24–26^, the effects of VR-mediated sensorimotor training across multiple sessions remain unknown. In summary, MI coupled with VR may boost sensorimotor learning and improve an individual’s ability to volitionally modulate EEG signals, thereby enhancing BCI classification to perform the virtual MI task. Mastery of MI BCI control is essential for advanced neurorehabilitation, particularly for individuals with SCI, who undergo cortical reorganisation and suffer significant declines in motor and sensory abilities following their injury^27^.

Here, we proposed a closed-loop sensorimotor training system deployed in VR to enhance the users’ modulation of EEG signals during lower-limb MI to improve BCI control (Fig. 1). The aim was to help individuals learn to regulate their sensorimotor activations and improve their ability to generate consistent and distinct MI patterns, thereby enhancing real-time BCI performance. To this end, we leveraged the principles of mirror therapy^28^ and designed a virtual environment to promote embodiment and motivation, allowing users to hopefully train effortlessly and regularly to perform a MI task on demand^29,30^. As opposed to simply using a mirror to show a limb, VR delivers multisensory real-time feedback (VR biofeedback) during repetitive lower-limb dynamic MI tasks, such as locomotion and cycling^31,32^. We hypothesised that using the proposed VR-mediated BCI training system over multiple sessions would result in neurophysiological adaptations, including enhanced neuroplasticity, cortical reorganisation, and functional connectivity, which would manifest as increasingly consistent and stable MI patterns in EEG signals. These adaptations are expected to strengthen neural circuits and promote more coherent oscillations in the alpha and beta bands, which are often associated with more efficient communication between sensorimotor cortical areas. Additionally, we anticipated that this training would improve neural efficiency by optimising sensorimotor pathways and activating the mirror neuron system, ultimately leading to improved BCI classification accuracy and greater user control over the MI virtual tasks. We first evaluated the developed system in five individuals with no sensorimotor impairment, *unimpaired*, and then, encouraged by our findings, we assessed an equivalent setup on four individuals with motor and sensory complete spinal cord injury. We showed that all participants learned to increase the control accuracy of BCI and improved their ability to produce distinct and stable EEG signals during relaxing and walking or cycling MI tasks. This is the first longitudinal study showing that individuals can learn to modulate sensorimotor activity associated with MI using an immersive VR-based BCI sensorimotor learning system. Additionally, this is the first longitudinal investigation into VR-mediated sensorimotor learning associated with lower-limb MI-BCI using consumer-grade hardware, incorporating both cued-online (MI initiated with VR auditory cues) and self-paced (MI initiated independently by the user without external cues) modes.

**Fig. 1:**
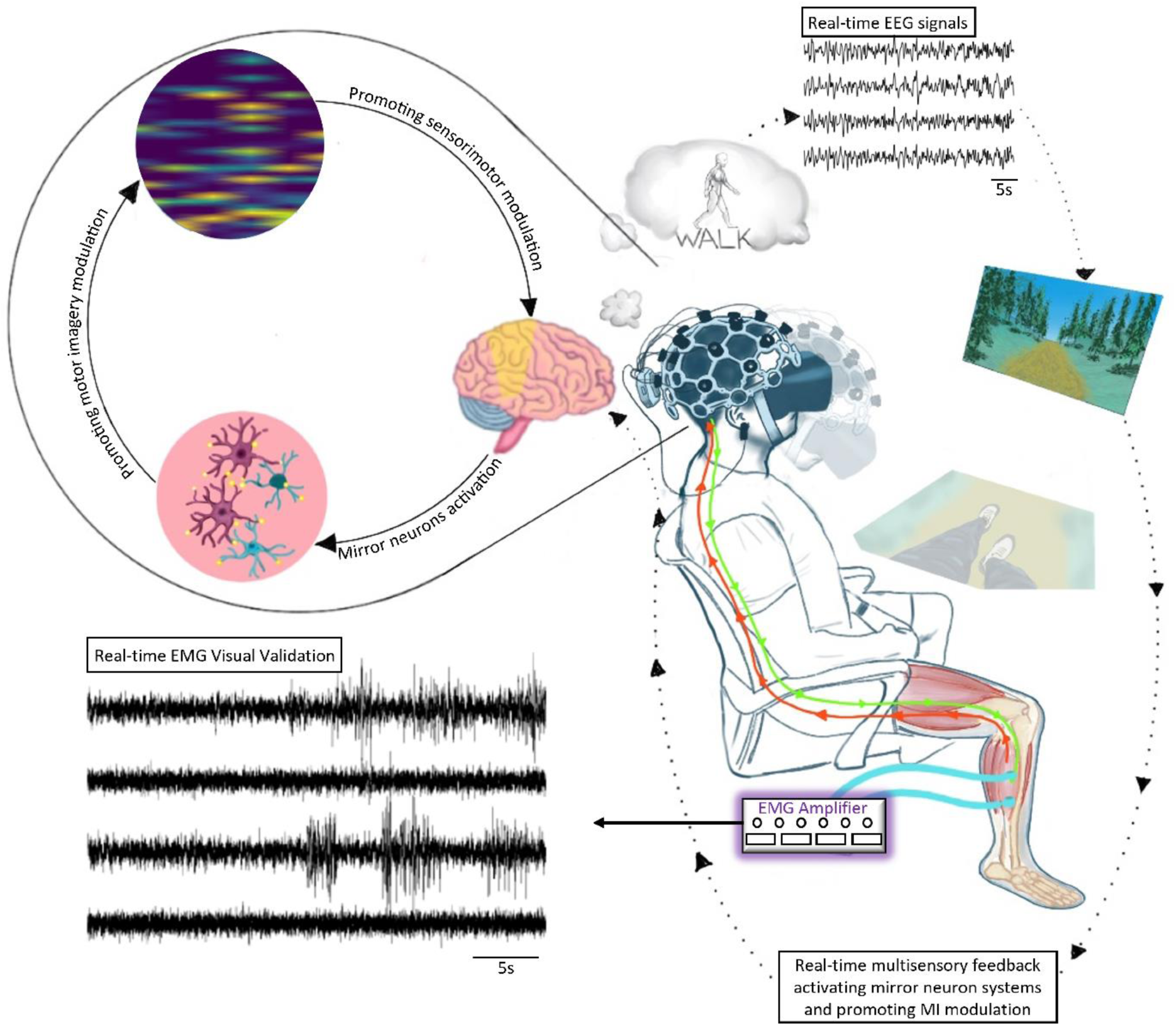
Design of the virtual reality mediated brain-computer interface sensorimotor training in a tailored immersive environment. Participants wore a 16-channel dry EEG system while immersed in a virtual pathed forest presented in an Oculus VR headset. EEG signals in response to different motor imagery (MI) tasks were decoded in real-time to control the movement of the avatar in the virtual environment. Training in an immersive virtual environment provided real-time visual and auditory sensory feedback possibly activating mirror neuron systems, resulting in stronger coordination of the primary sensorimotor cortex, and promoting neuromodulation of regular and rhythmically distinctive MI-EEG signals. Electromyography (EMG) signals were used to visually assess low-level background EMG activity in response to different MI tasks.

## Results

### Unimpaired individuals learned walking in VR

The training resulted in EEG patterns that were progressively more distinct and stable between the ‘relax’ and ‘walk’ MI classes (Fig. 2a-d). Specifically, the group average class discrimination (Fig. 2a) in the alpha band significantly increased with the number of training sessions (*r=*0.77, *p<*0.001), while no significant relationships were found for the beta bands (*r=*0.39, *p=*0.14). Significant negative linear relationships were observed between the test-train adaptation (TTA) and the number of training sessions (Fig. 2c) in both alpha (*r=*-0.70, *p<*0.005) and beta bands (*r=*-0.65, *p<*0.005). At the group level, comparing the first and last three sessions revealed a significant increase in class discrimination (Fig. 2b) and a decrease in TTA (Fig. 2d).

**Fig. 2:**
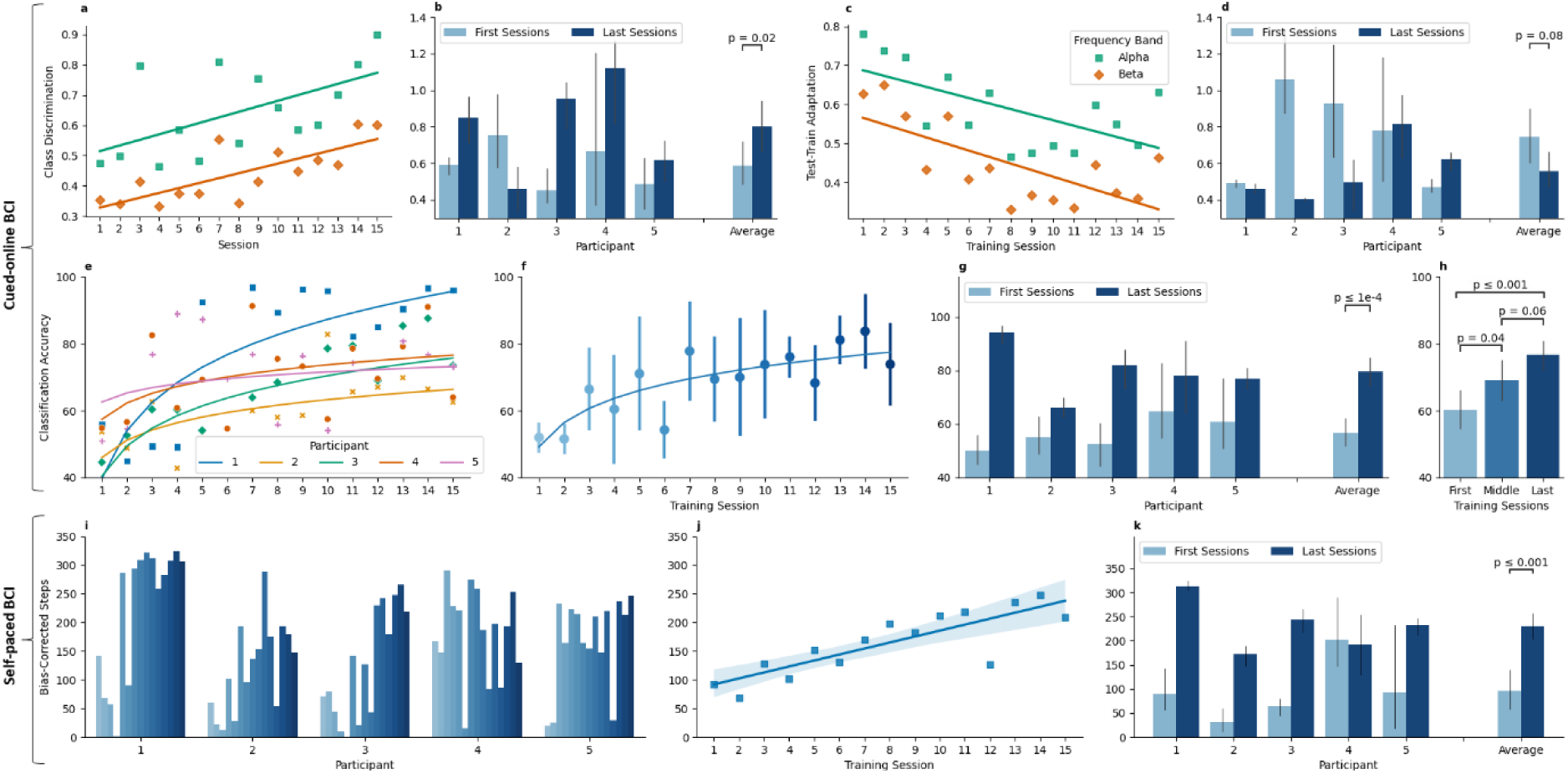
Virtual reality-mediated sensorimotor training improves BCI control and neuromodulation of sensorimotor rhythms at neuroimaging level for the unimpaired group. Training effect on the neuromodulation of sensorimotor rhythms as **a** class discrimination and **c** test-train adaptation (TTA) over the increasing number of training sessions in the alpha and beta EEG frequency bands for MI-BCI cued-online operation. Comparison of the first three and last three sessions’ class discrimination **b** and TTA **d** in the 4-30 Hz frequency band for all participants. **e** Classification accuracy, expressed as a percentage, and fitted curves (solid lines) for each participant in all fifteen sessions. Fitted curves are used to visualise the learning trend in controlling the BCI. **f** Group average classification accuracy, expressed as a percentage, for each session with vertical lines indicating the standard deviation. **g** Comparison of the first three and last three sessions’ classification accuracies for all participants. **h** Comparison of first, middle, and last five sessions’ group average classification accuracies. Real-time control of the self-paced BCI with **i** the steps taken are shown in the fifteen sessions by each participant, **j** the group average with a linear fit (solid line) showing increased learning with the number of sessions undertaken, and **k** in the first three and last three sessions.

For all unimpaired participants, their improved distinct and stable EEGs transferred to improved cued-online BCI classification accuracy for MI task performance with the number of training sessions undertaken (Fig. 2e-f). At the group level, the BCI classification accuracy significantly increased in the last three sessions compared to the first three (Fig. 2g, 57±10% vs. 80±11%, *p<*0.001), demonstrating progressive increments over the number of sessions (Fig. 2f and 2h). Across all participants the first, middle, and last five cued-online sessions each had significantly improved BCI classification accuracies (Fig. 2h), being the first (61±14%) vs. middle (69±16%); *p<*0.05, middle (69±16%) vs. last (77±11%); *p*=0.06; and first (61±14%) vs. last (77±11%), *p<*0.001).

During self-paced MI-BCI, the participants were well able to produce avatar walking in the virtual environment, and this improved with training. Generally, the number of steps increased with the number of training sessions (Fig. 2i), increasing linearly for the group average (*r=*0.84, p<0.001) (Fig. 2j). Except for one participant, all the participants successfully increased their number of steps from the first to the last three sessions (Fig. 2k, group average: 96±80 vs. 231±55, *p<*0.001).

None of the self-assessment metrics were found to be statistically significantly correlated with BCI classification accuracy for all participants (Supplementary Material Table 1). However, participant-specific predictors could be obtained.

### The developed system responded to lower-limb MI

The VR biofeedback enabled unimpaired participants to increase MI neuromodulation for real-time BCI classification, thereby enabling cued-online control in performing the MI task. To assess whether the user correctly evoked lower-limb MIs, electromyograms (EMGs) were measured bilaterally from the medial gastrocnemius and tibialis anterior muscles. Low-level background EMG activity increased following the ‘walk’ audio cue compared to the ‘relax’ audio cue, which was congruent with the EEG classification of the MI tasks by the BCI (Fig. 3 and supplementary video).

**Fig. 3:**
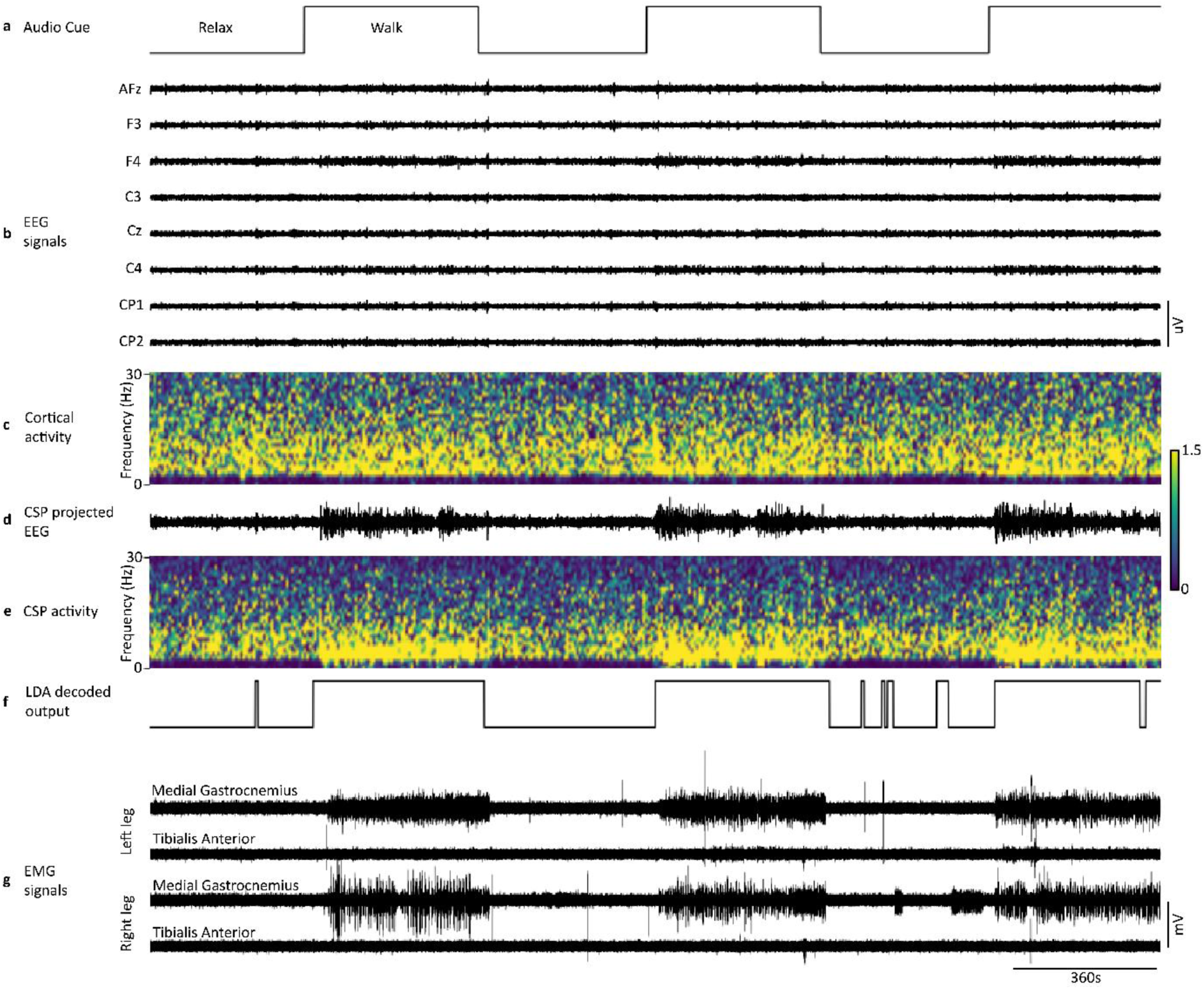
Representative data during real-time cued-online control of the BCI-commanded avatar. **a** Audio cue (“relax” or “walk”) instructing the participants to evoke the appropriate motor imagery (MI) task. **b** Electroencephalograms (EEG) for 8 representative channels. **c** Spectogram of the EEG signals recorded from the Cz channel. **d** The common spatial pattern (CSP) filter, of which parameters are calculated during the BCI calibration phase, was applied in real-time to all recorded EEG signals; here, the filtered Cz signal was reported, showing the increased signal variance between the “relax” and “walk” MI tasks. **e** Spectrogram of the CSP-filtered EEG from the Cz channel. **f** The output of the CSP filters was decoded via a linear discriminant analysis (LDA) to predict “relax” or “walk” MI; a few seconds delay can be observed between the audio cue and the decoded output, additionally, a few incorrect classifications can also be observed. **g** Bilateral electromyograms (EMG) from tibialis anterior and medial gastrocnemius were collected for a limited number of participants and sessions to visually assess the evoked MI corresponding to lower-limb activity. “Walk” MI corresponded with increased EMG background activity without muscle contraction (also see supplementary video).

### Individuals with SCI learned cycling in VR

All participants learned to produce more distinct EEG patterns with the training (Fig. 4a-d). They improved their class discrimination scores from the first to the last three sessions (Fig. 4e) except for one participant, whereas no difference was observed in TTA (Fig. 4f-j). During cued-online BCI, each participant with sensorimotor complete SCI improved their ability to control the BCI with an increasing number of training sessions undertaken (Fig. 4k, *r=*0.54, *p<*0.015). All individuals numerically improved their BCI classification accuracy from the first to the last three sessions (Fig. 4l, group average: 57±14% vs. 71±9%, *p<*0.01).

**Fig. 4:**
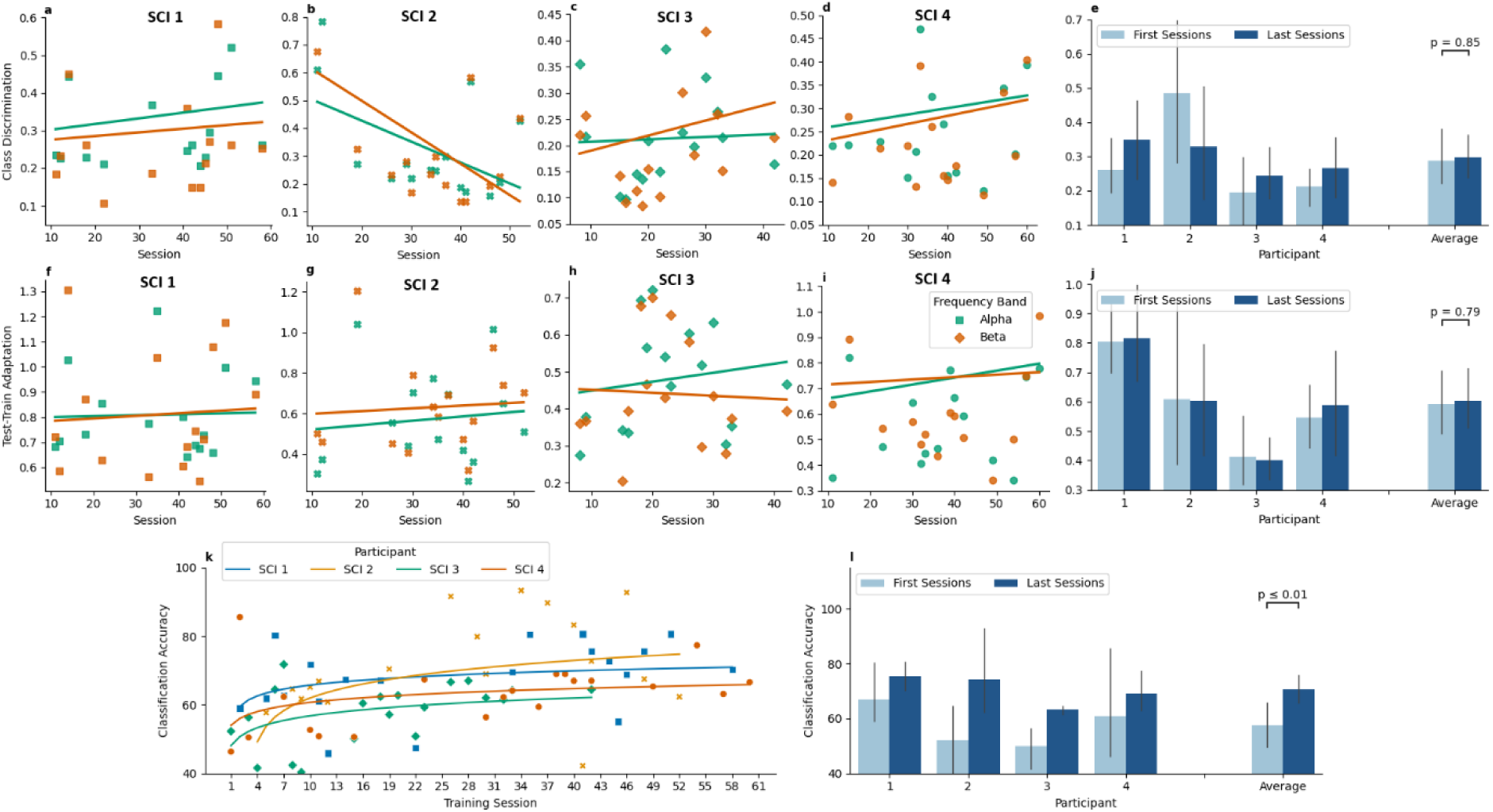
Real-time control of the BCI-commanded avatar for individuals with motor and sensory complete SCI. Only 20 training sessions were selected for each participant. The analysis did not include sessions with high noise or no cued-online data. Training effect on the neuromodulation of sensorimotor rhythms as **a-d** class discrimination and **f-i** test-train adaptation (TTA) over the increasing number of training sessions in the alpha and beta EEG frequency bands for MI-BCI cued-online operation. Comparison of the first three and last three sessions’ class discrimination **e** and TTA **j** in the 4-30 Hz frequency band for all participants. **k** Classification accuracy, expressed as a percentage, and fitted curves (solid lines) for each participant. **l** Comparison of the first three and last three session’s classification accuracies for all participants.

## Discussion

This study provides insights into the VR-mediated longitudinal sensorimotor training using consumer-grade dry EEG derived MI BCI system. The system enabled real-time control of virtual walking or cycling through both cued-online and self-paced modes, helping individuals enhance their ability to produce stable MI patterns. In agreement with our hypotheses, the VR mediated BCI training facilitated users to produce increasingly more consistent and stable underlying signatures of MI patterns in EEG signals during the lower-limb MI and virtual walking or cycling. This led to statistically significantly improved control (20% for unimpaired individuals and 14% for individuals with SCI) of the BCI-commanded avatar in the VR environment compared to baseline.

The lower-limb EMG activity showed that the real-time VR mediated BCI system correctly responded to walking MI (Fig. 3 and supplementary video). During the test performed on a single unimpaired participant, no bursts of EMG associated with muscle contraction were observed, meaning that the task was only imagined and not executed. Importantly, and consistent with previous reports^33,34^, the background EMG activity increased during walking compared to resting MI, indicating alignment between imagined movement and provided cue.

Improvements in the BCI classification accuracies alone do not necessarily suggest enhanced modulation of brain signals^35,36^. The purpose was to train individuals to enhance their abilities to control sensorimotor activations and produce stable MI patterns using the designed VR-mediated BCI system. Unimpaired participants learned to evoke more stable and distinct EEG patterns during MI as training progressed, which was revealed in two indices of EEG signal separability, class discrimination^35^, and TTA^36^. Class discrimination measures the stability and distinguishability of EEG signals within a class and between classes, respectively, over sensorimotor training sessions. Meanwhile, TTA assesses a user’s ability to consistently produce similar EEG signals for each class during calibration and cued-online operations within each training session. Learning to modulate EEG during MI is reflected in the gradual increase of class discriminability and/or a gradual decrease in TTA^35,36^. Our results showed a positive correlation between the average class discrimination and the number of training sessions (Fig. 2a-b); and a negative correlation between the group average TTA and session index (Fig. 2c-d), suggesting a learning effect. Interestingly, participant 1 of the unimpaired group exhibited no significant correlation for TTA (*r=*-0.15, *p* = 0.58), although achieving the highest BCI classification accuracy. Despite these general results, participant 1 learned to produce distinct EEG patterns for both classes (*r=*0.8, *p<*0.001) instead of adapting to the BCI classifier, indicating that individuals can improve their BCI performance either by producing distinct EEG responses or by adapting to the requirements of the classifier, as also reported in previous studies^36^. These considerations suggest that new approaches to assessing and studying BCI learning, namely, class discrimination and TTA, might be explored in the future.

Sustaining a MI for long periods is the key to intuitive and robust BCI control for efficient rehabilitation^12,13,37^. Unlike the conventional approach, which uses 3 to 6 seconds of MI tasks with rest periods, we designed the cued-online BCI to sustain MI tasks for a long time, 60 seconds, without any rest periods. VR biofeedback was provided to enhance the immersiveness, embodiment, and ease in sustaining MI for long time periods, which we hypothesised would help participants to improve self-paced BCI control with training. Furthermore, our BCI implementation enabled combining cued-online and self-paced modalities within the same session, a feature that has never been fully explored in longitudinal BCI training^13^. In general, acceptable self-paced BCI control requires training from a few sessions in a single day to many sessions over several days^16,35,38^, even requiring weeks for lower-limb MI-BCI-enabled rehabilitation^13^. Although our experiment was not designed to directly compare the combined effect of self-paced mode and real-time VR feedback to traditional approaches, like a fixation cross, static images, or otherwise, it might be speculated that the ability of the users to quickly learn to sustain MI and control the gating of the avatar walking or cycling (Fig. 2i-k) might have been facilitated by these technological advancements. Our results from cued-online and self-paced modalities are encouraging for the development of lower-limb BCIs where users need to maintain particular MIs for non-specific periods.

The Pearson correlation between BCI performance, being classification accuracies, and self-prediction by answers to a questionnaire for each participant was assessed using data from both the calibration and the cued-online phase (Supplementary Material). None of the self-predictors was found to be statistically significantly correlated with BCI performance across all participants. However, each participant had different self-predictors correlated with their classification accuracies. For instance, difficulty in evoking walking MI was found to be negatively correlated with BCI performance for two participants. Similarly, sleep, the physical state before the session, and engagement during the session were observed to be positively correlated with BCI performance for different participants. These results align with previous studies that investigated the relationship between the self-prediction of BCI performance with actual classification accuracies^39^, and further suggest that individual-specific metrics could be used as self-predictors of BCI control.

The goal of this study was to develop a BCI system that could be used in sensorimotor rehabilitation to train end-users to improve their BCI control^40–42^. Although advancements in BCIs for rehabilitation have significantly improved the quality of life for individuals recovering from strokes or (SCI)^5–9^, challenges remain with inconsistent BCI control^42^, which can disrupt effective feedback delivery. Furthermore, individuals with SCI undergo cortical reorganisation and experience declines in motor and sensory functions^27^, making it increasingly difficult for them to achieve efficient BCI control. Our VR-mediated BCI training system enabled individuals with SCI to enhance control of their avatar to perform various virtual tasks (Fig. 4), similar to what was observed in the unimpaired participants (Fig. 2). The rationale for improved learning and MI modulation using VR biofeedback was supported by recent studies showing that VR-based therapy resulted in stronger activations of the primary sensorimotor cortex than the conventional mirror therapy^43,44^. A plausible, but speculative, interpretation of our results is that our proposed VR-mediated BCI training might have improved learning through a combination of mirror neurons activation^45^, embodiment^30,31^, and multisensory feedback^31,32^. Furthermore, engagement and immersiveness via VR could have enhanced active participation and attention while performing the MI task, which has been shown to play an important role in promoting learning and plasticity^8,46,47^.

The adoption and acceptance of BCI technology by end-users and the clinical translation of BCI-enabled neurorehabilitation could be expedited by optimizing the technology to reflect end-users’ performance expectations and design priorities^42,48^. We addressed some of those priorities identified by the end-users by developing a system that included easy and quick setup, high classification accuracy, reliability, and consistency^40–42,48^, all of which pose considerable barriers to the use of BCI-enabled neurorehabilitation in clinics and home settings. Extensive setup and cleaning times for gel-based EEGs^12–14,36,49^ and expensive research-grade wireless EEGs^15^ are major hurdles preventing the widespread usage of MI-BCIs in clinical settings^50^. We selected a consumer-grade dry-electrodes EEG system that could be easily set up and is low-cost. Additionally, previous studies trained participants with upper-limb MIs that are comparatively easy to perform and decode, due to better access in recording cortical neural activity non-invasively, when compared with lower-limb tasks^51^. Furthermore, most of these studies employed a small number of training sessions^12–15^ and the performance enhancement might have only been indicative of transitory effects rather than consolidated user learning^49^. Weber et al.^52^ showed that more than ten sessions were required to attain good modulation of sensorimotor rhythms and subsequently obtain good predictions of participant performance. Indeed, naïve participants attained significant improvements in BCI control using our system (Fig. 3), establishing that these improvements were more persistent than what was previously reported^12–15^.

The significant improvements observed in BCI control, classification accuracy, and stable MI EEG signals during VR-mediated BCI training suggest neuroplasticity^53,54^, including cortical reorganisation^55^, enhanced functional connectivity^56,57^, and increased neural efficiency^58^. Repeated MI tasks within immersive VR environments likely promote long-term potentiation^59^, which strengthens neural circuits, improves synaptic efficiency, and facilitates coherent oscillations in alpha and beta bands—critical for motor control^60^. These synchronized oscillations likely facilitate more efficient and accurate decoding of motor intentions by the BCI system^61^. Additionally, the immersive multisensory feedback in VR supports cross-modal plasticity^62^ and could increase neural efficiency by optimizing sensorimotor pathways^58^, refining motor representations^62^, and activating the mirror neuron system^63^, resulting in more stable and distinct MI responses. Dopamine-mediated reward-based plasticity^64^ likely further supports these adaptations by reinforcing successful MI task performance. Collectively, these mechanisms underscore the potential of VR-mediated BCI training to drive neuroplasticity, optimize motor pathways, and enhance motor recovery, particularly for individuals with SCI.

This study has some limitations that should be considered. First, there was no control group to compare the learning effects of the proposed VR-BCI system with the conventional BCI system without VR. Recent studies, of less than three training sessions, showed that visual guidance via VR could help users to increase engagement and perform hand MI^21,24–26^. However, we showed that the VR-based training progressively improved distinct and stable EEGs with corresponding improvements in MI-BCI classification. Future studies should consider comparing the performance of BCIs with VR and no VR conditions across training sessions. Second, the experimental paradigm could be further optimized to enhance user engagement, especially the self-paced BCI. A possible improvement could be the introduction of multitasking and gamification to further engage and motivate the user^21^. The VR environment could also be improved by enhancing the sense of embodiment with the avatar. In this study, we used a generic avatar for all participants, but participant-specific modifications, like gender, could give users a more immersive experience^13^. Third, more advanced signal processing algorithms for feature extraction and classification algorithms could be used to improve the BCI performance^65^. Channel selection strategies could be utilized to select participant-specific EEG electrodes and further reduce the complexity of the system^15^.

In conclusion, we developed a VR-mediated consumer-grade BCI-controlled sensorimotor training system and showed improved avatar control during virtual walking and cycling. Both unimpaired individuals and individuals with SCI learned to modulate their EEG patterns during MI over several training sessions, with associated improvements in BCI classification accuracy. The results support the use of cued-online and self-paced VR biofeedback operations to enhance participant learning, showing encouragement for the development of low-cost BCI-enabled neurorehabilitation systems.

## Materials and Methods

### Unimpaired Participants

Seven right-handed unimpaired naïve participants (2 females, aged 24-40 years) participated in this study. Participants had no medical history that could have influenced the task, such as pregnancy, claustrophobia, drug addiction, or neurological disease. Each participant underwent 15 sessions of BCI training with each session taking place on a different day, with a maximum of three sessions per week, except for two participants who had longer intervals due to COVID-19 lockdowns and restrictions. Each participant’s subsequent sessions were set as early as their next availability to reduce the inter-session intervals. Two participants were excluded from the analyses because they did not complete all 15 sessions due to personal matters. All procedures and protocols were performed in accordance with the guidelines of the Declaration of Helsinki and have been approved by the Institutional Review Board of Griffith University (GU ref no: 2021/170). All participants provided their informed and signed consent prior to study participation.

### Experimental setup and BCI implementation

Each participant sat on a comfortable chair wearing an Ultracortex Mark IV EEG headset with cyton-daisy biosensing board (OpenBCI, USA), and VR headset (Oculus Rift S, Oculus, USA). The experiments consisted of two different MI tasks: relaxing and walking MI. Each session consisted of an offline calibration phase, and two user-controlled online operations, being cued-online BCI, and self-paced BCI. There was a 1-to-3 minutes break between each phase, based on user readiness; consequently, each session, including user preparation and data collection, lasted approximately one hour (Fig. 5a).

**Fig. 5:**
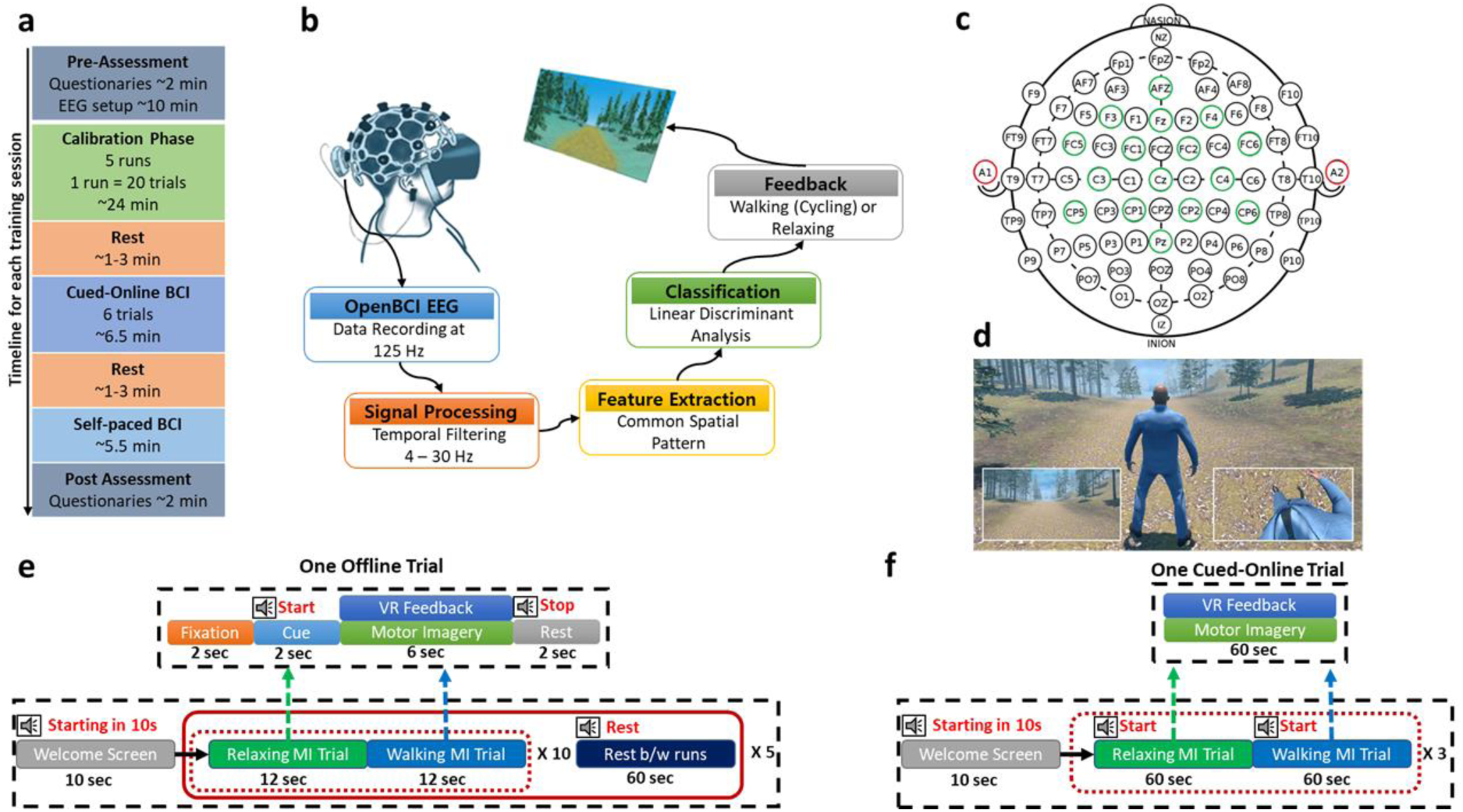
Schematic of the study. **a** Timeline for each training session. **b** BCI data collection and signal processing pipeline. Each session started with a calibration phase to obtain a participant-specific classification model followed by online BCI control (cued-online and self-paced BCI). **c** EEG electrode configuration (green circles) with reference and ground electrodes (red circles). **d** Virtual environment illustrating avatar within the simulated environment with inset images showing the first-person view, as seen by the user. **e** Experimental workflow for calibration phase. **f** Experiment workflow for cued-online phase.

The MI-BCI was implemented within the Robotic Operating System (ROS)^66^ to enable real-time EEG data acquisition and processing, as well as synchronisation with the VR environment (Fig. 5b). All EEG signals were pre-processed followed by features extraction via the common spatial pattern (CSP) method. A linear discriminant analysis (LDA) classifier was trained using calibration data to decode the users’ intentions for real-time BCI control. To assist calibration and BCI use, depending on the BCI classification, concurrent real-time multisensory VR feedback was provided to the user via relaxed standing or walking avatar. Finally, the progress of the participants in learning the BCI across the 15 training sessions was evaluated using three metrics: (i) classification accuracies, being the number of correct predictions divided by the total number of predictions in cued-online BCI-controlled VR walking, (ii) the number of steps in self-paced BCI-controlled VR walking, and (iii) ability to produce distinct and stable MIs evaluated using class discrimination and TTA. Details of all these steps are now presented.

### EEG Recordings

EEG signals were recorded at 125 Hz with a 3D printed headset fitted with 16 dry electrodes. The electrode placements for unimpaired participants were AFz, F3, Fz, F4, FC5, FC1, FC2, FC6, C3, Cz, C4, CP5, CP1, CP2, CP6, and Pz, whereas for individuals with SCI, the electrodes were placed at AFz, FC3, FC1, FCz, FC2, FC4, C3, C1, Cz, C4, C2, CP3, CP1, CPz, CP2, and CP4. The reference and ground electrodes were attached to the right and left earlobes, respectively (Fig. 5c). Electrodes were positioned according to a 10–20 system^67^ to cover the pre-motor, primary motor, and sensorimotor areas.

### Virtual Reality Environment

A virtual pathed forest (Fig. 5d), developed in Unity 3D (Unity Technologies, USA), was used to immerse the participants in a VR environment. A generic human avatar, obtained from the Unity asset store, was scaled to be proportional to the size of the VR environment and displayed in a first-person view. For the unimpaired group, the avatar was animated using a generic human walking animation obtained from Mixamo (Adobe, USA). For the SCI group, a tricycle was integrated into the VR environment, with cycling animation using Unity’s inbuilt physics system. The avatar moved or remained still according to the corresponding MI task, walking, cycling, or relaxing.

### Individuals with SCI

Four individuals with motor and sensory complete SCI (Table 1) participated in this study. All procedures and protocols were performed in accordance with the guidelines of the Declaration of Helsinki and have been approved by the Institutional Review Board of Griffith University (GU ref no: 2019/994). All participants provided their informed and signed consent prior to study participation. Each participant underwent multiple sessions of training, however, here we only reported 20 sessions and rejected noisy sessions after data inspection. Differently from the protocol used in unimpaired individuals, the VR environment was converted to visualize a cycling motor task, rather than walking (Fig. 6). Additionally, the EEG headset was modified to place electrodes over the motor and sensorimotor areas.

**Fig 6:**
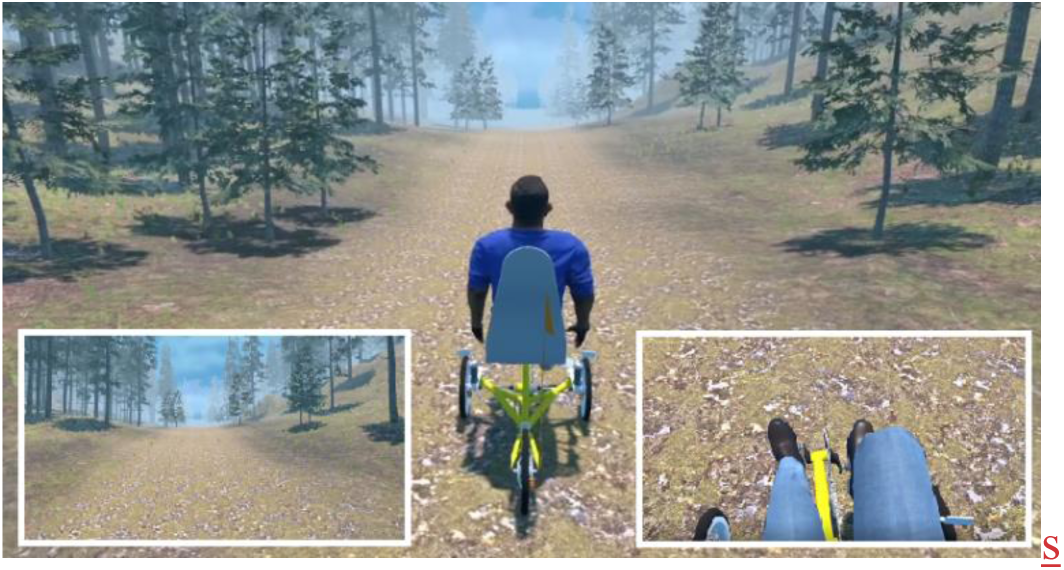
Virtual environment for the SCI group illustrating cycling avatar within the simulated environment with inset images showing the first-person view as seen by the user.

**Table 1:** Demographics of the individuals with motor and sensory complete spinal cord injury (SCI) and their American Spinal Injury Association Impairment Scale (AIS) classification level. ^68^.

| Participant | Age Range (years) | Gender | Type of lesion | Time since injury (months) |
| --- | --- | --- | --- | --- |
| SCI 1 | 35-40 | Male | C4, AIS A | 50 |
| SCI 2 | 45-50 | Male | T1, AIS A | 182 |
| SCI 3 | 40-45 | Male | C4, AIS B | 38 |
| SCI 4 | 35-40 | Male | C6, AIS A | 165 |

### Pre-processing

EEG data were filtered between 4–30 Hz using a zero-phase bandpass finite impulse response filter using the MNE library in Python^69^. In the offline calibration and cued-online operation phases, data epochs for relaxing and walking MI were automatically extracted according to task information coded within the ROS environment.

### Feature Extraction

CSP was employed for feature extraction of the two-class BCI; walking or cycling; or relaxing^70^. CSP decomposed the multichannel EEG data to generate a set of spatial filters that minimised the variance of one class while maximising the variance of the second class^65^. Thus, the discrimination between the two classes’ features was optimised by maximising the interclass variance and minimising the intraclass variance.

Mathematically, CSP works as follows. Let us assume *E*_*c*_^*k*^ represents the EEG data for trial *k* of class *c*, being, walking, cycling or relaxing. The dimensions of each *E*_*c*_^*k*^is *ch* × *s*, where *ch* is the number of channels and *s* is the number of samples per channel. Furthermore, let us assume that *E*_*R*,*W*_represents the EEG matrices for relaxing (*R*) and walking (*W*) MI classes, so the normalized spatial covariance for both classes are

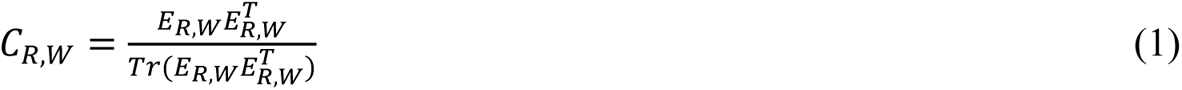

where *C*_*R*_and *C*_*W*_represent the covariance matrices for relaxing and walking classes, respectively, *T* is the transpose operator, and *Tr* is the trace of a matrix. The averaged covariance matrices *C̅*_*R*_and *C̅*_*W*_can be estimated by averaging *C*_*R*_and *C*_*W*_over all trials, respectively, with the composite spatial covariance matrix *C* calculated and diagonalized by

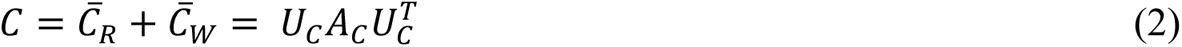

where *U*_*C*_ is the matrix of eigenvectors, and *A*_*C*_ is the diagonal matrix of eigenvalues. Therefore, the whitening transformation matrix *P* can be defined by *U*_*C*_ and *A*_*C*_ as

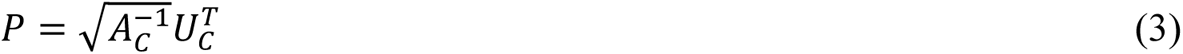

which equalizes the variances in the space spanned by *U*_*C*_ being, all eigenvalues of *PCP*^*T*^are equal to one. *C̅*_*R*_ and *C̅*_*W*_are transformed as

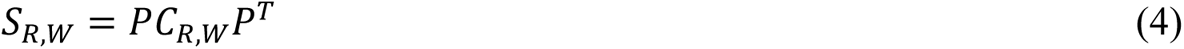

then *S*_*R*_ and *S*_*W*_ share the common eigenvectors, which is,

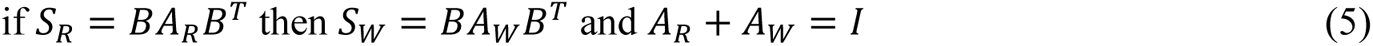

where *B* is the matrix of eigenvectors and *I* is the identity matrix. Since the sum of two corresponding eigenvalues is always one, the eigenvectors with the largest eigenvalue for *S*_*R*_ has the smallest eigenvalues for *S*_*W*_and vice versa. This property makes the eigenvectors *B* useful for the classification of the two distributions. The projection of the whitened EEG onto the first and last eigenvectors in B (the eigenvectors corresponding to the largest *A*_*R*_and *A*_*W*_) will give feature vectors that are optimal for discriminating two populations of EEG. Thus, the spatial filters *W* can be obtained as

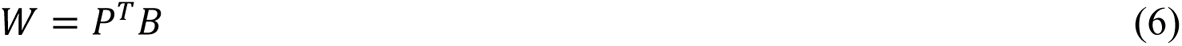

Then the features *Z*_*R*_ and *Z*_*W*_ can be obtained after the application of *W* to filter the original EEG signals as

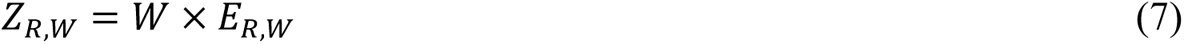

In general, the first and last *m* filters of the CSP projection result in the most discriminative features between the two classes. Therefore, each MI trial was represented by selecting 2*m* rows. As a result, *v*_*R*_and *v*_*W*_were chosen as the feature vectors of relaxing and walking MI classes, with the definition as

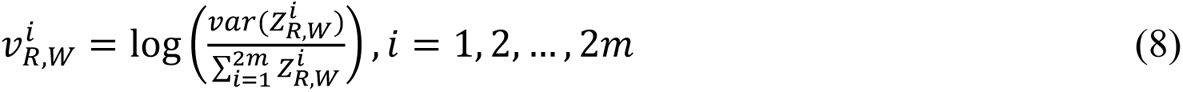

In this study, *m* was chosen as 2.

### Classification Algorithm

LDA was selected as classification algorithm because it has low computational demands and provides good results for two-class classification, thus making it ideal for our real-time and online BCI^71^. LDA projects feature vectors extracted using CSP from one specific dimensional space to another with the conditions of maximizing the between-class differences and minimizing the within-class scattering.

Thus, LDA finds a hyperplane that maximizes the distance between the means of the projected CSP features in (8) for the two classes, relaxation and walking^71^. For this two-class classification problem, a discriminant function has a decision hyperplane defined by

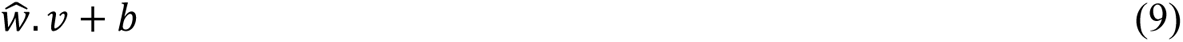

where *w̑* is a vector of classification weights, *v* are the vectors of features being, 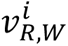 in (8), and *b* is the bias term. The weight vector *w*^ is calculated through

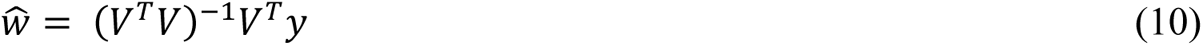

where *V* is the matrix containing feature vectors for each class and *y* is the vector with class labels (0 for relaxation and 1 for walking).

### Experimental Workflows

#### Calibration Phase

Each session began with a calibration phase (Fig. 5e), wherein participants underwent alternating periods of relaxing and walking MI with corresponding VR feedback of the imagined task while their EEG data were collected. Calibration consisted of 5 runs with each run consisting of 10 trials for each MI task (100 trials). Each trial lasted for twelve seconds and included a fixation cross (t = 0 – 2s) indicating the commencement of a new trial, audio cues (t = 2 – 4s) prompting participant to perform the desired MI task (t = 4 – 10s), and a final audio cue indicating the completion of the trial (t = 10 – 12s). A break of one minute was included after the completion of each run. Participants were asked to avoid unnecessary eye blinking and body movements, especially during the imagination period. A familiarization session on day 1 was performed before the actual data acquisition to allow participants to fully understand the experimental protocol so that they could perform MI tasks to the best of their ability.

#### Cued-online Phase

After calibrating the classification model, the cued-online phase (Fig. 5f) was conducted to evaluate the performance of each participant. This phase started with a welcome screen (t = 0 – 10s). The cued-online phase involved 3 trials (60s each) for each MI task, with an audio cue to prompt the participant to perform the desired MI task for 60s, with real-time VR biofeedback consistent with the decoded MI command. Unlike the calibration phase, there were no fixation, cues, or rest periods between the trials. Overall, the cued-online experiment lasted six minutes and ten seconds.

#### Self-paced Online Phase

In the self-paced online phase, participants were instructed to take as many steps as possible in the VR environment within five minutes, which had the goal of enhancing the users’ abilities to maintain a particular imagination task for long periods.

##### Performance Metrics

Classification accuracy is a criterion for evaluating the performance of a participant in controlling the BCI and producing stable MI responses. True positive (TP, walking MI performed by the participant and correctly predicted by classifier), false positive (FP, relaxing MI performed by the participant but walking MI predicted by classifier), true negative (TN, relaxing MI performed by the participant and correctly predicted by classifier), and false negative (FN, relaxing MI performed by the participant but walking MI predicted by classifier) were used to calculate the classification accuracy in each session as

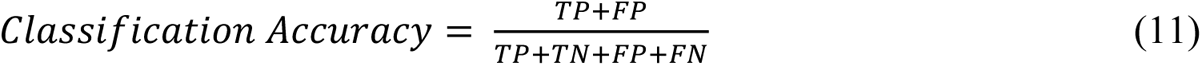

In addition, logarithmic curve fitting, *a* + *b log*(*x*) where *x* is the classification accuracy of a session, and *a* and *b* are the fitting coefficients, was performed to visualise the trend in classification accuracies for each participant across 15 training sessions. Curve fitting was performed using the SciPy module in Python. Improvements in individual MI classification, defined as individual command scores, were calculated as the number of correct predictions divided by the total number of trials for that MI class (see supplementary material).

Besides classification accuracy, class discrimination and TTA were used to evaluate participant progress and to study the changes in the characteristics of EEG patterns at neuroimaging level. Class discrimination defines as the stability and distinctness of the EEG patterns produced by the user, independent of any classifier^35^. Class discrimination (*classDis*) for a two-class problem was calculated as

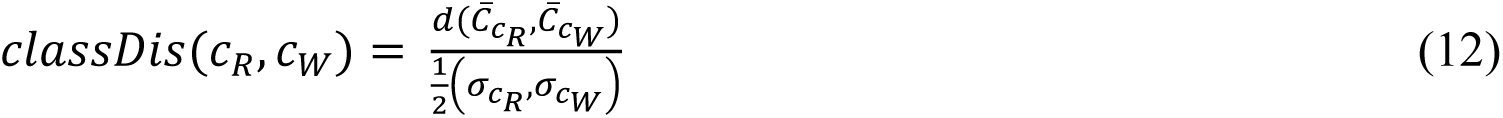

where the numerator computes the Riemannian distance between class means and the denominator is the summation of average squared distances around the Karcher mean of each class. Overall, increased class discrimination over the sessions would suggest the enhancement in the user’s ability to produce distinct and stable EEG responses for each class.

TTA examines the user’s adaptation to the MI-BCI system and classifier. TTA was used to estimate the similarity/dissimilarity between calibration and online (cued-online and self-paced) phases^36^

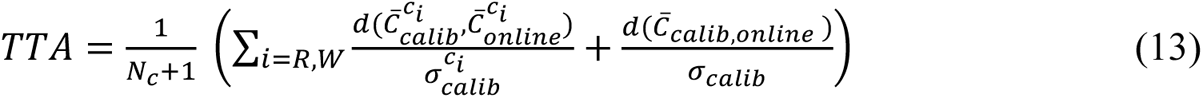

where *N*_c_ is the number of classes (*N* = 2), 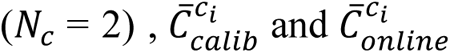 are the Riemannian means of class *c*_*i*_ (walking and relaxing) in calibration and *online* phases, respectively, 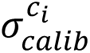 is the standard deviation of class *c*_*i*_ in calibration data, 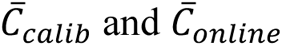 denote the global means, and *σ*_*calib*_ is the global standard deviation of all calibration and *online* samples. In contrast to class discrimination, a decreasing TTA over the training sessions would indicate more adaptation of the *online* phase data compared to the calibration phase data. In other words, this would mean that the participant is learning to produce EEG patterns increasingly more similar to what the classifier expects for each class.

The primary outcome measure for the self-paced BCI phase was the number of steps taken within five minutes in each training session; however, this metric could be affected by the bias of the trained model. To normalise the measured number of steps across training sessions and participants, the trained model was assumed to be biassed towards any of the two MI classes, which would therefore affect outcomes, being the number of steps. We calculated the corrected number of steps (*Steps*_*Corr*_) by reducing the biassedness in the recorded number of steps (*Steps*_*Rec*_) as

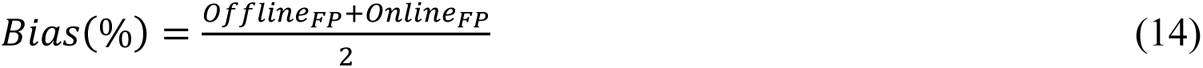

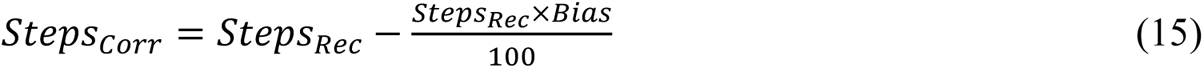

where *Offline*_*FP*_and *Online*_*FP*_are the false positive rates in calibration and cued-online phases, respectively. We used FP rates to reduce biasedness towards the walking MI.

#### Statistical Analysis

Pearson correlation between classification accuracies and session index was used to show the learning effects of the training with the VR-mediated BCI system and to quantify if any of the questionnaire’s subjective metrics could be used to predict the performance of an individual. Additionally, the BCI performance in the first three and last three sessions was compared and tested for significant differences at the 95% confidence interval using Wilcoxon nonparametric rank-sum tests^49^.

#### Validation using EMG Signals

Surface EMG signals were recorded at 2048 Hz throughout the BCI session with four electrodes using an OT Bioelettronica Quattrocento EMG amplifier. The electrodes were bilaterally placed at the medial gastrocnemius and tibialis anterior muscles. Skin preparation and electrode placement were performed according to SENIAM guidelines. For real-time demonstration (Supplementary video), raw EMG signals were high-pass filtered at 10 Hz.

## Data Availability

All data produced in the present study including supplementary video are available upon reasonable request to the corresponding authors.

## Acknowledgments

This work was supported by the Queensland Motor Accident Insurance Commission (MAIC), Australia, as part of the BioSpine research project. For this study, Y.D.T. was supported by the Australia-Harvard Fellowship.

## Authors Contributions

MMNM wrote the first draft and was involved in conceptualization, study design, software development, data collection with unimpaired and SCI groups, and data analysis.

DP provided critical feedback and was involved in the study design.

KM was involved in VR environment development and data collection with the SCI group.

EJ was involved in the EEG headset design used for data collection with the SCI group and preparation of Figure 1.

EL was involved in data collection with the SCI group.

AQ was involved in software development.

CC was involved in the initial stages of software development and BCI setup.

FR was involved in data collection and EMG data analysis.

YDT was involved in revising the first draft.

DGL and CP supervised the overall study, provided critical feedback, and were involved in conceptualization, study design, reviewing, and revising the paper.

