## Supplementary Material for "Virtual reality mediated brain-computer interface training improves sensorimotor neuromodulation in unimpaired and post spinal cord injury individuals"

Target Journal: Nature Communications

* Corresponding authors

Prof Yang D. Teng,

Dr Claudio Pizzolato,

Keywords: Brain-computer interface (BCI), spinal cord injury, electroencephalogram (EEG), consumer-grade dry-electrodes, motor imagery, sensorimotor learning, mirror therapy, virtual reality, lower-limb, common spatial pattern (CSP), linear discriminant analysis (LDA)

### *Questionnaire*

A questionnaire was used to assess the relationship between an individual’s subjective assessment of their MI-BCI performance and their quantitative classification accuracy. To-this-end, before and after each session all participants completed a questionnaire that consisted of 15 questions on a 10-point scale with 1 being easy/bad/low and 10 being hard/good/high, except for the questions about sleep and drink which were answered in hours. The questions were

Before session

i. How many hours did you sleep last night?
ii. When do you have your last drink (caffeine/alcohol)?
iii. How difficult is it for you to perform relaxing MI?
iv. How difficult is it for you to perform walking MI?
v. What is your physical state?
vi. What is your mental state?
vii. What is your fatigue level?
viii. What is your motivation level for this session?

After session:

i. How do you rate the protocol and VR environment? (First session only)
ii. What was your engagement level during the session?
iii. Did you get bored?
iv. What was your concentration level during the session?
v. What is your physical state?
vi. What is your mental state?
vii. What is your fatigue level?

### Results

**Table 1:**  Mean (standard deviation) questionnaire-based self-assessment metrics for each participant with the Pearson correlation with the classification accuracies in calibration and cued-online operation (*p<*0.05).

| Subject | Correlation  with classification accuracies | Metrics | | | | | | | | | | | | | |
| --- | --- | --- | --- | --- | --- | --- | --- | --- | --- | --- | --- | --- | --- | --- | --- |
|  |  | **Pre-experiment** | | | | | | | | **Post-experiment** | | | | | |
|  |  | **Sleep** | **Drink** | **Relaxing MI** | **Walking MI** | **Physical State** | **Mental State** | **Fatigue Level** | **Motivation** | **Engagement** | **Boredom** | **Concentration** | **Physical State** | **Mental State** | **Fatigue Level** |
| 1 |  | 8.00 (0.0) | 9.87 (4.0) | 5.00 (0.0) | 5.67 (0.9) | 9.00 (0.53) | 9.27 (0.45) | 1.07 (0.59) | 10.00 (0.0) | 8.60 (1.84) | 6.13 (0.99) | 8.13 (1.24) | 7.60 (0.73) | 7.93 (0.26) | 6.93 (1.03) |
|  | Calibration |  |  |  | **-0.82** |  | **-0.68** |  |  | **0.96** |  | **0.65** | **0.64** |  |  |
|  | Cued-online |  |  |  | **-0.90** | **-0.56** | **-0.84** |  |  | **0.93** |  | **0.62** |  |  |  |
| 2 |  | 8.13 (0.9) | 23.7 (10) | 1.87 (0.6) | 5.33 (1.1) | 8.27 (0.7) | 7.80 (0.6) | 2.07 (0.5) | 7.93 (0.6) | 7.27 (1.5) | 2.80 (1.4) | 5.00 (0.9) | 8.13 (0.6) | 8.33 (0.5) | 5.27 (1.3) |
|  | Calibration | **0.55** |  |  | **-0.68** | **-0.64** |  |  |  |  |  |  | **-0.68** |  | **-0.79** |
|  | Cued-online | **0.63** |  |  | **-0.71** | **-0.74** |  |  |  |  |  |  | **-0.69** |  | **-0.71** |
| 3 |  | 7.59 (0.5) | 10.7 (9.5) | 1.53 (0.8) | 4.47 (0.9) | 9.27 (0.8) | 7.87 (0.5) | 3.13 (0.5) | 7.4 (0.6) | 7.00 (0.6) | 6.93 (0.6) | 6.80 (0.8) | 8.07 (0.3) | 7.13 (0.3) | 5.13 (1.7) |
|  | Calibration | **0.70** | **-0.52** |  |  |  |  |  |  |  |  |  |  |  | **-0.87** |
|  | Cued-online | **0.65** |  |  |  | **-0.63** |  |  |  |  |  |  |  | **0.54** | **-0.60** |
| 4 |  | 6.33 (1.0) | 12.53 (8) | 5.93 (0.9) | 3.20 (1.4) | 8.00 (1.1) | 7.33 (0.7) | 3.07 (1.4) | 5.53 (1.3) | 5.40 (0.5) | 4.20 (0.8) | 5.73 (1.1) | 7.80 (1.2) | 6.07 (0.8) | 4.67 (1.4) |
|  | Calibration |  |  |  |  |  |  |  |  |  |  |  |  |  |  |
|  | Cued-online |  |  |  |  |  |  |  |  |  |  |  |  |  |  |
| 5 |  | 7.67 (0.7) | 2.17 (1.2) | 4.2 (1.5) | 1.0 (0.4) | 7.86 (0.7) | 7.6 (1.1) | 3.8 (2.0) | 7.0 (2.1) | 6.2 (2.1) | 5.67 (2.3) | 6.13 (2.1) | 6.73 (1.8) | 5.86 (2.2) | 7.0 (2.6) |
|  | Calibration |  |  |  |  |  |  |  | **-0.56** | **0.57** |  |  |  |  |  |
|  | Cued-online |  |  |  |  |  |  |  |  | **0.72** |  |  |  |  |  |
| Average |  | 7.54 (0.9) | 11.8 (9.9) | 3.7 (1.9) | 3.96 (1.9) | 8.5 (0.9) | 7.98 (0.9) | 2.61 (1.5) | 7.56 (1.8) | 6.9 (1.8) | 5.15 (1.9) | 6.36 (1.6) | 7.68 (1.2) | 7.08 (1.4) | 5.73 (1.9) |
|  | Calibration |  |  | **0.26** |  |  | **0.24** | **-0.27** | **0.32** | **0.55** |  | **0.36** |  | **0.24** |  |
|  | Cued-online |  |  | **0.33** |  | **-0.30** |  |  |  | **0.45** |  | **0.44** |  |  |  |

### Individual Command Scores

In calibration phase, there were significant linear increases in the true positive (TP) (*r=*0.7, *p<*0.004) and true negative (TN) scores (*r=*0.81, *p<*0.001) over the training sessions for both relaxing and walking MI (Figure S1A, C left panels). There were also linearly decreased false positive (FP) (*r=*-0.81, *p<*0.001) and false negative (FN) (*r=*-0.7, *p<*0.004) error rates over the training sessions (Figure S1B, D left panels). From the first three sessions to the last three sessions, each individual showed greater TP and TN command accuracies, and lower FP and FN command errors, with the only exception of TN and FP for participant 4 (Figure S1A-D right panels). Overall, there were significantly increased group average command accuracies from the first three sessions to the last three sessions (TP: 64%±6 vs. 80%±10, *p<*0.01; TN: 61%±9 vs. 81%±12, *p<*0. 01) and significantly decreased command errors (FP: 39%±9 vs. 20%±12, p<0.01; FN: 36%±6 vs. 20%±10, p<0.01).

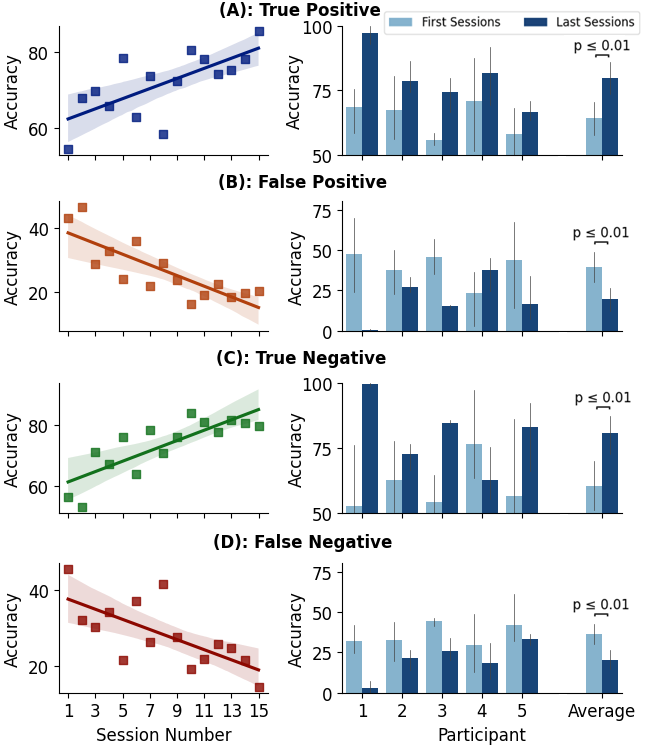

**Figure S1:** Calibration phase group average command scores with the linear fit (solid line) (left panels), and comparison of individual command scores in the first three and last three sessions for each participant and group average (right panels).

In the cued-online phase, command accuracies linearly increased (Figure S2A, C left panels: TP: *r=*0.47, *p* = 0.07, TN: *r=*0.66, *p<*0.007) while command errors linearly decreased (Figure S2B, D left panels: FP: *r=*-0.47, *p<*0.07, TN: *r=*-0.66, *p<*0.007) with the number of training sessions. Except for participant 4, from the first to the last three sessions' command accuracies and error rates improved (Figure S2A-D right panels). From the first to the last three sessions, the group averages revealed significantly increased TN (52%±17 vs. 81%±15, *p<*0.03) and significantly decreased FP (48%±18 vs. 19%±11, *p<*0.03). However, no significant differences for TP (62%±18 vs. 78%±11, *p<*0.33) and FN (38%±17 vs. 22%±15, *p<*0.33) were found, although, both these metrics showed numerically better results in the last three compared to the first three sessions.

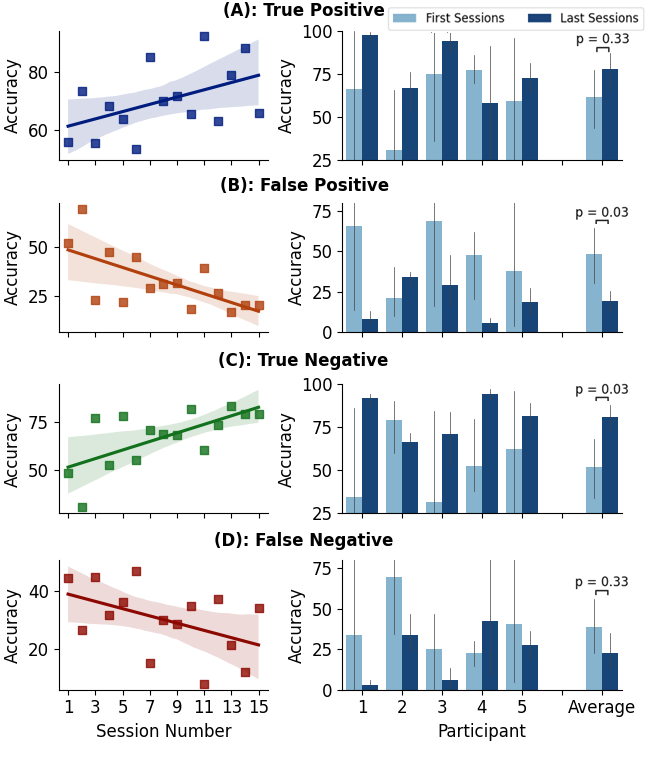

**Figure S2:** Cued-online phase group average command accuracies with the linear fit (solid line) (left panels) and comparison of individual command scores in the first three and last three sessions for each participant and group average (right panels).

Improvements in individual command accuracy demonstrated participant learning and adaptation to the system using the proposed VR-mediated consumer-grade BCI-controlled sensorimotor training. The average command accuracy had a positive learning effect on TP and TN (Figure S1-S2), and all participants were able to significantly reduce command errors by 15-20% for both walking and relaxing (Figures S1-S2) from the first to the last sessions.
